# State Xylazine Scheduling and Changes in Xylazine and Medetomidine Reports in the U.S. Illicit Drug Supply: A Quasi-Experimental Study

**DOI:** 10.64898/2026.05.12.26353052

**Authors:** David T. Zhu, Sehun Oh

## Abstract

**Background:** Xylazine and medetomidine are veterinary sedatives increasingly detected as adulterants in the U.S. illicit drug supply. In response, several states have scheduled xylazine. Whether these policies are associated with subsequent changes in xylazine and medetomidine detections remains unclear.

**Methods:** We conducted a state-level, semiannual, serial cross-sectional study using National Forensic Laboratory Information System (NFLIS) data from 1999 to 2025. The primary outcomes were xylazine and medetomidine reports per 100,000 NFLIS drug reports. We used staggered difference-in-differences event-study models to estimate changes in report rates after xylazine scheduling. Sensitivity analyses excluded Florida and expanded the treatment definition to include states that criminalized xylazine without formal scheduling. Falsification analyses examined steroid and antidepressant reports as negative-control outcomes.

**Results:** NFLIS recorded 101,987 xylazine reports and 12,085 medetomidine reports. Xylazine scheduling was not associated with a significant change in xylazine report rates (ATT, 2,872.29 per 100,000; 95% CI, −2,024.63 to 7,769.21; *p*=.250). In contrast, xylazine scheduling was associated with a significant increase in medetomidine report rates (ATT, 1,536.51 per 100,000; 95% CI, 211.14 to 2,861.88; *p*=.023). Sensitivity analyses produced similar findings. Negative-control outcomes showed no significant changes.

**Conclusions:** State xylazine scheduling was associated with increases in medetomidine reports but no significant change in xylazine reports. These findings suggest that scheduling may be followed by changes in adulterant composition rather than reductions in overall α2-adrenergic agonist involvement. Our study highlights the importance of monitoring the unintended effects of xylazine scheduling and supporting continued investment in public health surveillance, drug checking, and harm reduction services.

## INTRODUCTION

Xylazine and medetomidine are α2-adrenergic receptor agonists approved exclusively for veterinary medicine as sedatives, analgesics, and muscle relaxants (D’Orazio et al., 2023; Cano et al., 2024; Palamar & Krotulski, 2024; Zhu et al., 2024). In recent years, both substances have been increasingly identified in the illicit drug supply in the United States (U.S.) (Zhu et al., 2024). Xylazine and medetomidine are most often detected in combination with illicitly manufactured fentanyl (IMF) (Friedman et al., 2022; Zhu et al., 2023; Palamar & Krotulski, 2024; Walton et al., 2025; Zhu & Palamar, 2025; Vohra et al., 2026). Xylazine-IMF overdose deaths have risen sharply, increasing from 99 deaths in 2018 to 6,020 deaths in 2023, corresponding to crude mortality rates of 0.03 to 1.80 per 100,000 population (Zhu & Cano, 2024).

In addition to overdose risk, xylazine and medetomidine can cause profound sedation, which may leave people who use drugs (PWUD) more vulnerable to theft, assault, and other forms of harm (Zhu, 2023). Xylazine has also been associated with necrotic skin ulcers that may require specialized wound care and, in severe cases, amputation (D’Orazio et al., 2023; Lutz et al., 2025). Medetomidine withdrawal can produce severe and potentially life-threatening hemodynamic and cardiovascular instability if abruptly discontinued (Philadelphia Department of Public Health, 2025; Zimmerman et al., 2026). Given their rapid spread across U.S. illicit drug markets and distinct toxidromes, xylazine and medetomidine have created new challenges for clinicians, harm reduction programs, and public health agencies.

Federal and state policymakers have responded to the emergence of xylazine and medetomidine adulteration through several regulatory and public health approaches. At the federal level, one of the earliest responses came from the White House Office of National Drug Control Policy, which designated xylazine-IMF mixtures as an “emerging drug threat” in April 2023 and issued a national response plan involving the Emerging Threats Committee and other key stakeholders in July 2023 (Zhu et al., 2023). Congress also introduced the Combating Illicit Xylazine Act in February 2025, which would federally schedule xylazine; however, the bill has not yet passed (Congress, 2025).

At the state level, several legislators have enacted laws that schedule xylazine and medetomidine as controlled substances or criminalize their possession, manufacture, and distribution (further details are presented in Table 1 and will be described more fully in the Methods). State public health agencies have also expanded the distribution of harm reduction resources, including naloxone, given the frequent co-adulteration of xylazine and medetomidine with IMF; test strips for xylazine, medetomidine, and IMF; confirmatory drug testing, such as liquid chromatography-mass spectrometry (LC-MS); and substance use education for PWUD about the distinct risks posed by these emerging adulterants (Quijano et al., 2023; Zhu, 2023; Palamar & Krotulski, 2024; Vickers-Smith et al., 2025; Crews, 2025; Zhu & Palamar, 2025).

**Table 1.** State Xylazine Scheduling and Criminalization Policies Through December 2025.

| State | Regulation Type<sup>a,b</sup> | Additional Details |
| --- | --- | --- |
| Delaware | Formal scheduling | Temporarily classified as a Schedule III controlled substance on June 2, 2023, and permanently classified (Senate Bill 189) on November 29, 2023. |
| Florida | Formal scheduling | Florida Statute 893.03, Chapter 2016-105: Permanently classified as a Schedule I controlled substance on July 1, 2016. |
| Nebraska | Formal scheduling | Legislative Bill 72: Permanently classified as a Schedule III controlled substance on February 25, 2025. |
| Ohio | Formal scheduling | Temporarily classified as a Schedule III controlled substance on March 29, 2023, and permanently classified (Rule 4729:9-1-03) on September 28, 2023. |
| Pennsylvania | Formal scheduling | Temporarily classified as a Schedule III controlled substance on June 3, 2023, and permanently classified (House Bill 1661) on May 15, 2024. |
| Rhode Island | Formal scheduling | House Bill 5922: Permanently classified as a Schedule V controlled substance on June 26, 2023. |
| South Carolina | Formal scheduling | House Bill 4617: Permanently classified as a Schedule III controlled substance on May 20, 2024. |
| South Dakota | Formal scheduling | Codified Laws 34-20B-20.2: Permanently classified as a Schedule III controlled substance on February 12, 2024. |
| West Virginia | Formal scheduling | Senate Bill 546: Permanently classified as a Schedule IV controlled substance on June 8, 2023. |
| Arizona | Criminalization | House Bill 2045: Added xylazine to the statutory definition of “dangerous drug” on April 8, 2024, thereby subjecting non-exempt possession, use, sale, manufacturing, transportation, and administration to Arizona’s dangerous-drug criminal penalties. |
| Indiana | Criminalization | House Bill 1203: Created criminal penalties for the non-exempt possession and distribution of xylazine, |
|  |  | effective March 11, 2024. |
| Louisiana | Criminalization | <u>House Bill 645</u>: Created criminal penalties for the unlawful production, manufacturing, distribution, or possession of xylazine, effective August 1, 2023. |
| Tennessee | Criminalization | <u>House Bill 1242</u>: Created criminal penalties for the non-exempt possession, manufacturing, delivery, or sale of xylazine, effective July 1, 2023. |
| Virginia | Criminalization | <u>House Bill 1187</u>: Created criminal penalties for the non-exempt manufacture, sale, distribution, or possession of xylazine intended for human consumption, effective July 1, 2024. |

<sup>a</sup>Formal scheduling refers to the state designation of xylazine as a controlled substance.
<sup>b</sup>Criminalization refers to state regulations that establish criminal penalties for non-exempt possession, manufacture, or distribution of xylazine (with narrow exceptions for veterinary use), without designating xylazine as a controlled substance.

Medetomidine may have emerged, at least in part, as a substitute adulterant after xylazine scheduling, as both drugs are α2-adrenergic receptor agonists with similar psychoactive effects, including profound sedation (Sugarman et al., 2024; Crews, 2025; Zhu et al., 2025). Prior descriptive and observational work has suggested that in some states, including Pennsylvania, xylazine scheduling laws temporally coincided with increases in medetomidine detections (Hochstatter et al., 2025; Zhu et al., 2025). Reports from state and local public health agencies are consistent with this pattern. The Philadelphia Department of Public Health has described “the introduction of medetomidine and concomitant decrease of xylazine in Philadelphia’s drug supply” (Puleo et al., 2025). Similarly, the Maryland Department of Health noted that “the emergence of medetomidine in Maryland coincides with a reduction in xylazine that followed the scheduling of xylazine in Pennsylvania and Virginia” (Maryland Department of Health, 2025).

Despite these observations, no study has examined whether xylazine scheduling may have had a potentially causal relationship with subsequent changes in xylazine and medetomidine detections. In this study, we leverage a quasi-experimental design to evaluate how state xylazine scheduling actions were associated with changes in xylazine and medetomidine reports in the Drug Enforcement Administration’s (DEA’s) National Forensic Laboratory Information System (NFLIS). We hypothesized that, compared with states that did not regulate xylazine, states that enacted xylazine scheduling would then experience lower rates of xylazine reports and higher rates of medetomidine reports.

## METHODS

### Study Design and Data Sources

In this serial cross-sectional study, we obtained data from NFLIS to examine changes in xylazine and medetomidine reports after implementation of state xylazine scheduling policies in the U.S. (Drug Enforcement Administration, 2025). NFLIS collects drug identification results from 284 participating federal, state, and local forensic laboratories across all U.S. states (Drug Enforcement Administration, 2023). We excluded Massachusetts because xylazine is effectively covered under the state’s longstanding Schedule VI framework since 1971 (General Court of the Commonwealth of Massachusetts, n.d.; McClure & Pless, 2023), making the timing of policy exposure less relevant to the recent state-level scheduling wave.

We obtained state-level data for semiannual periods between 1999 and 2025, with each calendar year divided into January to June (first half-year period [H1]) and July to December (H2). Semiannual periods were used because they are the most granular time units available in the public NFLIS database. NFLIS classifies medetomidine and dexmedetomidine separately: we included only medetomidine because it is not approved for human use and represents an illicit drug adulterant, whereas dexmedetomidine is approved for clinical use, including intensive care unit and procedural sedation (Barends et al., 2017; Lewis et al., 2022).

### Exposure

The primary exposure was formal state scheduling of xylazine as a controlled substance. To identify relevant laws, we first reviewed the March 2024 state xylazine legislation summary from the Legislative Analysis and Public Policy Association (Legislative Analysis and Public Policy Association, 2024), as well as a content analysis of xylazine policy responses (Sugarman et al., 2024). Because our study period extended through 2025, we supplemented these sources with a manual review of LegiScan, state legislative websites, and news articles to identify more recent laws and verify their effective dates. Table 1 presents all state xylazine laws identified through December 2025.

Nine states have formally scheduled xylazine as a controlled substance: Delaware (Delaware Department of State, 2023; Delaware General Assembly, 2023), Florida (McClure & Pless, 2023; Florida Legislature, 2025), Nebraska (Nebraska Legislature, 2025), Ohio (Office of the Governor of Ohio, 2023; Ohio Laws and Administrative Rules, 2023), Pennsylvania (Commonwealth of Pennsylvania, 2023; LegiScan, 2024a), Rhode Island (Rhode Island General Assembly, 2023; LegiScan, 2023), South Carolina (South Carolina General Assembly, 2024), South Dakota (South Dakota Legislature, 2024), and West Virginia (West Virginia Legislature, 2023; Campbell, 2023).

Five additional states enacted laws that impose criminal penalties for the non-exempt possession, manufacture, sale, or distribution of xylazine while preserving narrow exemptions for veterinary use: Arizona (LegiScan, 2024b), Indiana (LegiScan, 2024c), Louisiana (Louisiana State Legislature, 2023), Tennessee (Tennessee General Assembly, 2023), and Virginia (Virginia Legislative Information System, 2024).

### Outcome

The primary outcomes were xylazine and medetomidine reports in each state-semiannual period. Because NFLIS reporting volume differs across states and over time (Cano et al., 2023), we obtained the total number of NFLIS drug reports for each state-semiannual period and used this value as the denominator. Consistent with prior methodologies (Pitts et al., 2023), we calculated xylazine and medetomidine report rates per 100,000 total NFLIS reports (hereinafter “per 100,000” for brevity).

### Data Analysis

In the descriptive analysis, we calculated mean xylazine and medetomidine report rates per 100,000 for states that formally scheduled xylazine (the treatment group) and for states that did not (the comparison group). Massachusetts and the five states that enacted criminal penalties without formally scheduling xylazine were excluded from the primary analysis.

The quasi-experimental approach used a difference-in-differences (DiD) analysis.

Because states enacted scheduling at different times, a traditional two-period DiD approach was not appropriate, as it does not allow for variation in treatment timing. We therefore estimated *staggered* DiD event-study models using the Callaway and Sant’Anna estimator (Callaway & Sant’Anna, 2021), which compares treated states with not-yet-treated and never-treated states during the same periods. Models were estimated across the five semiannual periods before and after each state’s xylazine scheduling date. Standard errors were clustered at the state level.

The models estimated the average treatment effect on the treated (ATT), defined as the average change in xylazine or medetomidine report rates among states that scheduled xylazine relative to the change that would have been expected in those same states in the absence of scheduling. The ATT represents the average effect across the entire post-scheduling period. We also estimated event-study coefficients, which represent effects for each individual semiannual period relative to scheduling, allowing us to examine whether changes emerged, strengthened, or attenuated over time.

Models adjusted for lagged xylazine burden, operationalized as the mean xylazine report rate per 100,000 during the two semiannual periods immediately preceding each state’s scheduling date. This adjustment mitigates potential policy endogeneity, as states with higher pre-policy xylazine burden may have been more likely to enact scheduling laws. By accounting for recent pre-policy xylazine burden, the models aimed to better isolate changes attributable to scheduling rather than to pre-existing increases in xylazine reports.

We assessed the parallel trends assumption, which assumes that treated and comparison states would have experienced similar trends in xylazine and medetomidine report rates in the absence of scheduling. We assessed this assumption in three ways. First, we visually inspected event-study coefficients for evidence of pre-policy divergence between treated and comparison states. Second, we examined whether individual pre-policy event-study coefficients differed significantly from zero. Third, we formally conducted a Wald test assessing whether all pre-policy event-study coefficients were jointly equal to zero.

We then conducted two sensitivity analyses. First, we repeated the analysis after excluding Florida because it scheduled xylazine in July 2016, substantially earlier than the next scheduling action in Ohio (March 2023). Second, we expanded the treatment group definition to include both the nine scheduling states and the five states that criminalized xylazine without formally scheduling it.

Finally, we conducted falsification analyses using steroid and antidepressant report rates as negative-control outcomes. These substances were selected because they are captured in the NFLIS database but should not be directly affected by xylazine scheduling. Evidence of post-policy changes in these unrelated drug classes would suggest that the main findings may reflect broader shifts in forensic testing, reporting, enforcement, or drug-market conditions. Conversely, null findings would support the interpretation that observed changes are more specific to xylazine scheduling.

The Virginia Commonwealth University Institutional Review Board deemed this study exempt from review because it used publicly available, deidentified data and did not constitute human subjects research. Analyses were performed using Stata version 19.5 (StataCorp LLC).

## RESULTS

### Descriptive Trends

NFLIS recorded 101,987 xylazine reports between 1999 and 2025. The vast majority of xylazine reports occurred in the most recent years of the study period, with 95,234 reports (93.38%) occurring between 2021 and 2025 (eTables 1–2). Medetomidine first appeared later, with 12,085 total reports, all occurring between 2021 and 2025.

Figure 1, eTable 3, and eFigures 1–2 show that states that ultimately scheduled xylazine already had higher xylazine report rates than comparison states before scheduling took effect. Among treated states, mean xylazine report rates increased from 940.57 per 100,000 at event time T_−5_ (five semiannual periods before scheduling) to 4,335.91 per 100,000 at T_−1_ (the semiannual period immediately before scheduling). Rates continued to rise after scheduling, peaking at 12,563.91 per 100,000 at T_2_ (two semiannual periods after scheduling), but this increase was not sustained. By T_5_, xylazine report rates roughly halved to 6,601.55 per 100,000. Comparison states showed a similar upward trend at lower levels, rising from 778.99 per 100,000 at T_−5_ to 1,988.06 per 100,000 at T_−1_, then peaking at 3,105.69 per 100,000 at T_2_.

**Figure 1.**
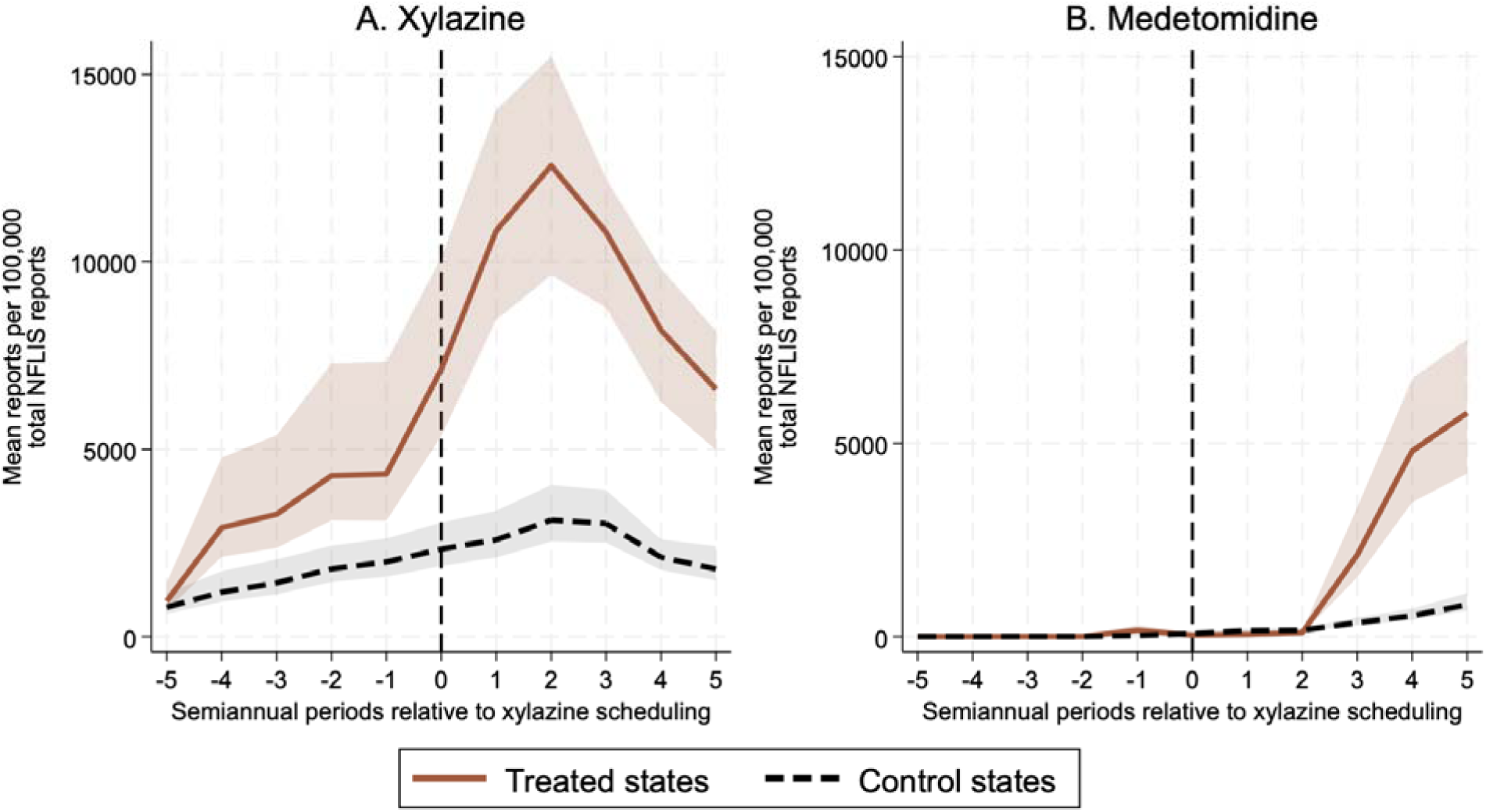
Changes in Mean Xylazine and Medetomidine Report Rates Relative to Index Semiannual Period of Xylazine Scheduling Caption. Line graph of mean xylazine and medetomidine reports per 100,000 state-level drug reports in the National Forensic Laboratory Information System (NFLIS). Shaded areas represen 95% CIs. The solid brown line represents states that scheduled xylazine as a controlled substance during the study period. The dashed black line represents states that did not schedule xylazine. Massachusetts was excluded because xylazine is covered under its longstanding Schedule VI framework. Arizona, Indiana, Louisiana, Tennessee, and Virginia were excluded because they enacted regulations criminalizing xylazine but did not formally schedule it. NFLIS data were obtained on May 7, 2026.

Medetomidine report rates were relatively low in both groups before xylazine scheduling. Among treated states, medetomidine was virtually not detected at T_−5_, but then increased to 160.65 per 100,000 at T_−1_. After scheduling, rates initially changed little, increasing from 37.57 per 100,000 at T_0_ to 100.96 per 100,000 at T_2_. However, in the later post-policy periods, medetomidine report rates increased sharply, reaching 5,783.23 per 100,000 at T_5_. Comparison states also had rising medetomidine report rates, but the increase was much smaller, from 74.83 per 100,000 at T_0_ to 823.21 per 100,000 at T_5_.

### Parallel Trends Assumption

Figures 2–3 and eFigures 3–4 show no clear visual evidence of differential pre-policy trends across the primary, sensitivity, or falsification models. eTable 4 shows that none of the individual pre-policy event-study coefficients were significant in any model. eTable 5 further shows that Wald tests, evaluating whether all pre-policy event-study coefficients were jointly equal to zero, had *p*>.05 for every model. Together, these checks support the parallel trends assumption and the internal validity of the DiD modeling approach.

**Figure 2.**
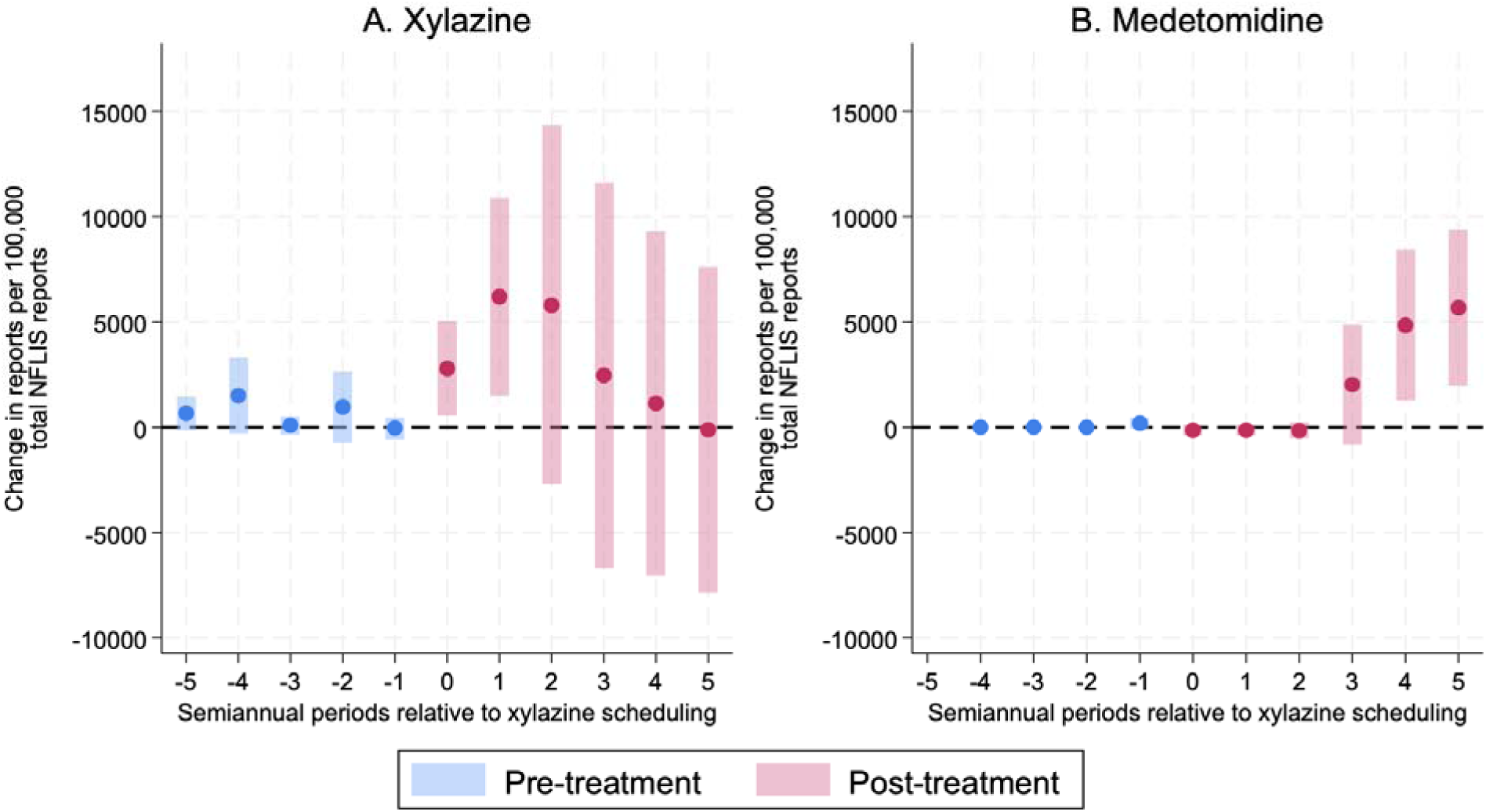
Event-Study Coefficients of Changes in Xylazine and Medetomidine Report Rates Following State Xylazine Scheduling Caption. Event-study coefficients of changes in xylazine and medetomidine reports per 100,000 state-level drug reports in the National Forensic Laboratory Information System (NFLIS) following state xylazine scheduling. Points represent estimated treatment effects and shaded bars represent 95% CIs. Blue estimates denote pre-treatment periods and pink estimates denote post-treatment periods. Five semiannual periods before and after scheduling were examined. Massachusetts was excluded because xylazine is covered under its longstanding Schedule VI framework. Arizona, Indiana, Louisiana, Tennessee, and Virginia were excluded because they enacted regulations criminalizing xylazine but did not formally schedule it. NFLIS data were obtained on May 7, 2026.

**Figure 3.**
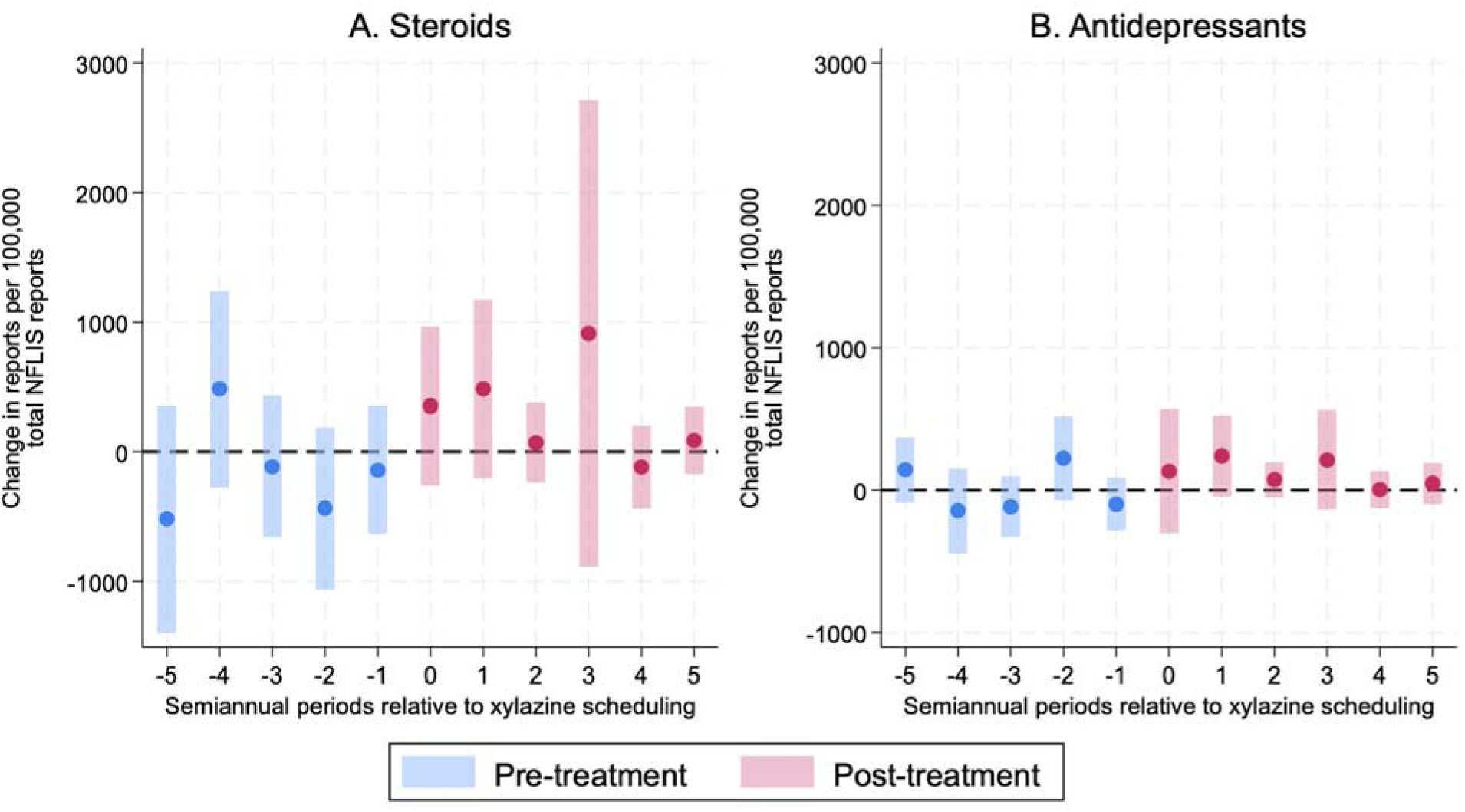
Fslsification Test Event-Study Coefficients of Changes in Steroid and Antidepressant Report Rates Following State Xylazine Scheduling Caption. Event-study coefficients of changes in steroid and antidepressant reports per 100,000 state-level drug reports in the National Forensic Laboratory Information System (NFLIS) following state xylazine scheduling. Points represent estimated treatment effects and shaded bars represent 95% CIs. Blue estimates denote pre-treatment periods and pink estimates denote post-treatment periods. Five semiannual periods before and after scheduling were examined. Massachusetts was excluded because xylazine is covered under its longstanding Schedule VI framework. Arizona, Indiana, Louisiana, Tennessee, and Virginia were excluded because they enacted regulations criminalizing xylazine but did not formally schedule it. NFLIS data were obtained on May 7, 2026.

### Primary DiD and Event Study Analyses

Figure 2 and eTable 4 show that xylazine scheduling was not associated with a significant overall change in xylazine report rates in the primary DiD analysis (ATT, 2,872.29 per 100,000; 95% CI, −2,024.63 to 7,769.21; *p*=.250). Event-study coefficients indicate a short-term increase immediately after scheduling at T_0_ (2,783.16 per 100,000; 95% CI, 543.75 to 5,022.57; *p*=.015) and T_1_ (6,194.35 per 100,000; 95% CI, 1,505.69 to 10,883.01; *p*=.010). After that point, the event-study coefficients declined and were no longer significant, suggesting that post-scheduling increase in xylazine reports tended to be short-lived.

In contrast, xylazine scheduling was associated with a significant overall increase in medetomidine report rates (ATT, 1,536.51 per 100,000; 95% CI, 211.14 to 2,861.88; *p*=.023). Early post-policy estimates were not significant, but the later estimates were large and significant at T_4_ (4,839.59 per 100,000; 95% CI, 1,273.79 to 8,405.40; *p*=.008) and T_5_ (5,677.21 per 100,000; 95% CI, 1,979.81 to 9,374.61; *p*=.003). Thus, unlike xylazine, which showed no significant overall change and only a short-lived early increase, medetomidine increased significantly after scheduling, with the strongest changes emerging in later post-policy periods.

### Sensitivity Analyses and Falsification Tests

Sensitivity analyses showed the same overall pattern. eFigure 3 and eTable 4 show that, after excluding Florida, the association betweem xylazine scheduling and xylazine report rates remained non-significant overall (ATT, 3,668.18 per 100,000; 95% CI, −3,175.77 to 10,512.14; *p*=.293). Medetomidine report rates remained significantly higher after scheduling, with an overall estimate slightly larger than in the primary analysis (ATT, 1,762.35 per 100,000; 95% CI, 333.49 to 3,191.20; *p*=.016). As in the primary analysis, increases in medetomidine report rates were concentrated in the later post-policy periods, at T_4_ (4,839.59 per 100,000; 95% CI, 1,273.79 to 8,405.40; *p*=.008) and T_5_ (5,677.21 per 100,000; 95% CI, 1,979.81 to 9,374.61; *p*=.003).

eFigure 4 and eTable 4 show similar results when the treatment definition was expanded to also include states that criminalized xylazine without formally scheduling it. The overall association between xylazine scheduling and xylazine report rates remained non-significant (ATT, 2,359.09 per 100,000; 95% CI, −1,793.17 to 6,511.35; *p*=.265). Medetomidine report rates again showed a significant overall increase following xylazine scheduling, although the estimate was slightly smaller than in the primary analysis (ATT, 1,253.87 per 100,000; 95% CI, 134.89 to 2,372.86; *p*=.028). The event-study coefficients were significant for medetomidine at T_4_ (4,839.59 per 100,000; 95% CI, 1,245.39 to 8,433.80; *p*=.008) and T_5_ (5,677.21 per 100,000; 95% CI, 1,909.84 to 9,444.57; *p*=.003).

Finally, Figure 3 and eTable 4 show that falsification analyses did not identify significant overall changes in steroid report rates (ATT, 138.34 per 100,000; 95% CI, −329.40 to 606.09; *p*=.562) or antidepressant report rates (ATT, 157.79 per 100,000; 95% CI, −4.78 to 320.37; *p*=.057). Most post-policy event-study coefficients for these negative-control outcomes were also not significant. These negative-control findings suggest that the medetomidine increase was unlikely to be explained by broader changes (e.g., NFLIS reporting, enforcement activity, forensic testing, or unrelated drug-market conditions), supporting the interpretation that the main findings were more specific to the effects of xylazine scheduling.

## DISCUSSION

To our knowledge, this is the first study to leverage a quasi-experimental design to evaluate whether state xylazine scheduling was associated with subsequent changes in xylazine and medetomidine report rates in NFLIS. We found that xylazine scheduling was associated with a significant overall increase in medetomidine reports, with the largest increases arising roughly one year after scheduling. In contrast, xylazine scheduling was not associated with a significant overall change in xylazine reports during the post-policy period. These findings are timely given rapidly evolving trends in xylazine- and medetomidine-involved overdoses and as state legislators and policymakers continue to consider whether scheduling and criminalization are appropriate responses to emerging adulterants in the U.S. illicit drug supply.

The most important takeaway from our analysis is that xylazine scheduling was followed by a substantial increase in medetomidine reports. This finding is consistent with our substitution hypothesis and with reports from state and local public health agencies suggesting that medetomidine began appearing as xylazine declined in some drug markets (Puleo et al., 2025; Maryland Department of Health, 2025). It is also consistent with what would be expected in an illicit drug market responding to a targeted regulatory shock. If scheduling made xylazine harder to obtain, easier to prosecute, or more costly to distribute, manufacturers and distributors may have had incentives to identify another sedating adulterant that could serve a similar role in fentanyl-adulterated products. Medetomidine may have been an attractive substitute because it is pharmacologically related to xylazine, has sedative α2-adrenergic agonist effects, and has higher potency, which may allow it to be transported or mixed in smaller quantities and evade detection (Sugarman et al., 2024; Crews, 2025; Zhu et al., 2025).

These findings should be interpreted within the broader context of how illicit drug markets have historically adapted to governmental interdictions. For example, the DEA’s 2018 federal class-wide scheduling of fentanyl-related substances was followed by declines in some fentanyl analogs, including carfentanil (Jalal & Burke, 2021). However, the synthetic opioid market continued to evolve, and this regulatory pressure may have contributed to the subsequent emergence of nitazene analogs (Roberts et al., 2022; Zhu et al., 2025). Within-class substitution patterns may have occurred within nitazenes themselves, as scheduling of one analog, such as isotonitazene, appears to have temporally coincided with the rapid rise of newer analogs, including metonitazene and protonitazene (Roberts et al., 2022; Zhu et al., 2025). Within this context, the rise of medetomidine after xylazine scheduling is concerning but ultimately not surprising: scheduling one α2-adrenergic agonist (xylazine) may shift the supply toward a related and more potent alternative (medetomidine) rather than reducing sedative adulteration overall.

Another major finding from our study is that xylazine scheduling was associated with no significant overall change in xylazine reports. This was inconsistent with our hypothesis that xylazine detections would decrease after scheduling. In fact, the event-study coefficients initially increased at T_0_ and T_1_, suggesting that xylazine reports briefly rose after scheduling before later declining and becoming non-significant. One likely explanation is policy endogeneity. States such as Pennsylvania, which were among the earliest epicenters of the U.S. xylazine overdose crisis (Friedman et al., 2022; Zhu, 2023; Cano et al., 2024), may have moved to schedule xylazine precisely because it was already rising rapidly in their illicit drug supply. Although our models adjusted for lagged xylazine burden to mitigate this concern, residual confounding may remain if scheduling decisions were influenced by sharp local increases, media attention, public health alerts, or law enforcement priorities that were not fully captured by recent pre-policy rates that were adjusted for (Quijano et al., 2023; Sugarman et al., 2024).

A second explanation is that scheduling may have changed detection and reporting behavior. Once xylazine was scheduled, law enforcement agencies, forensic laboratories, and public health officials may have become more attentive to it (Pitts et al., 2023). This could increase xylazine detection or reporting activity in the short term, even if the underlying supply was beginning to stabilize or decline. A third potential reason is the relatively short post-policy period. The later event-study coefficients for xylazine appeared to follow a downward trajectory and became non-significant. Because most state scheduling decisions examined in our study occurred recently in 2023 and 2024, our event-study window included only five semiannual periods of follow-up. That may be too short to observe whether xylazine declines are sustained. Continued monitoring is therefore needed to determine whether xylazine reports continue to fall, whether medetomidine keeps rising, or whether the market shifts again toward newer substitutes.

### Areas for Future Research

Our findings also point to several priorities for future research and monitoring. Clinical and epidemiological studies should continue to track the spread of xylazine, medetomidine, and other α2-adrenergic agonists, as well as their patterns of co-adulteration and substitution. Future work should also examine whether the observed post-scheduling patterns in xylazine and medetomidine drug reports extend to complementary data sources, including overdose mortality, toxicology, emergency department visits, and wound care encounters. Finally, qualitative and mixed-methods research with PWUD, drug checking programs, forensic scientists, clinicians, and harm reduction organizations will also be valuable to explain how medetomidine enters local markets, to what extent people are aware of its presence, and how its clinical effects differ from xylazine in real-world settings.

### Policy Implications

Our findings ultimately raise concerns about the effectiveness of xylazine scheduling and provide early evidence that these policies may unintentionally accelerate substitution toward other α2-adrenergic receptor agonist adulterants such as medetomidine. This is especially important as states begin considering similar regulatory approaches for medetomidine, with South Dakota recently becoming the first state to schedule medetomidine (as a Schedule III compound) in March 2026 (LegiScan, 2026).

Thus, further efforts are needed to prioritize public health surveillance, drug checking, clinical preparedness, and harm reduction resources. This could include expanding access to xylazine, medetomidine, and IMF test strips; increasing the availability of confirmatory testing such as LC-MS; funding harm reduction programs to counsel PWUD about prolonged sedation, wound risk, and withdrawal symptoms; and preparing clinicians to manage xylazine-related necrotic wounds, medetomidine withdrawal-related cardiovascular instability, and clinical features (Quijano et al., 2023; Zhu, 2023; Palamar & Krotulski, 2024; Vickers-Smith et al., 2025; Crews, 2025; Zhu & Palamar, 2025). Naloxone distribution remains essential because these substances are often co-adulterated with fentanyl (Zhu, 2023). Nevertheless, naloxone alone cannot address the full toxidrome of α2-adrenergic agonists, emphasizing the need to prioritize the development of specific reversal agents (Mullins & Seger, 2025).

### Limitations

Several limitations warrant consideration. First, NFLIS records drug reports from participating forensic laboratories and may not capture all illicit drug supply and seizure events in the U.S. (Pitts et al., 2023). Testing practices may vary across states and over time, and increased awareness of xylazine or medetomidine could influence detection. We partially addressed this concern by standardizing xylazine and medetomidine reports as rates using state-semiannual period level counts of total NFLIS reports as the denominator, but this approach does not fully account for differences in testing practices. Second, the analysis used semiannual data because this was the most granular publicly available time unit, which limited temporal precision around exact policy implementation dates. Third, although we adjusted for lagged xylazine burden and conducted several tests to assess the parallel trends assumption, unmeasured time-varying confounding remains possible, particularly because states may have scheduled xylazine in response to pre-existing upward trajectories in xylazine’s prevalence and enforcement priorities. Finally, most xylazine scheduling policies were relatively recent, leaving a relatively short post-policy window for evaluating longer-term effects.

## CONCLUSION

Overall, our study found that state xylazine scheduling was associated with a significant overall increase in medetomidine report rates but no significant overall change in xylazine report rates. These findings suggest that scheduling may shift the composition of the illicit drug supply rather than reduce the overall burden of α2-adrenergic receptor agonist adulteration. As xylazine, medetomidine, and other novel adulterants continue to spread across the U.S. and contribute to overdoses and other health risks, our findings provide evidence that state legislators should consider moving beyond reactive scheduling. Public health agencies should prioritize real-time epidemiological surveillance, reduce barriers to accessing xylazine and medetomidine test strips and drug checking, and strengthen clinical responses to ever-evolving illicit drug markets.

## Data Availability

All data produced in the present work are contained in the manuscript.

## Notes

### Competing Interest Statement

The authors have declared no competing interest.

### Funding Statement

This study did not receive any funding.

### Author Declarations

The Virginia Commonwealth University Institutional Review Board deemed this study exempt from review because it used publicly available, deidentified data and did not constitute human subjects research.

