## Supplementary Tables and Figures for "State Xylazine Scheduling and Changes in Xylazine and Medetomidine Reports in the U.S. Illicit Drug Supply: A Quasi-Experimental Study"

**eTable 1. Number of Xylazine Reports in NFLIS by Semiannual Period, 1999–2025**

| **State** | **Semiannual Period** | | | | | | | | | | | |
| --- | --- | --- | --- | --- | --- | --- | --- | --- | --- | --- | --- | --- |
|  | **1999 H1** | **1999 H2** | **2000 H1** | **2000 H2** | **2001 H1** | **2001 H2** | **2002 H1** | **2002 H2** | **2003 H1** | **2003 H2** | **2004 H1** | **2004 H2** |
| Alabama |  |  |  |  |  |  |  |  |  |  |  |  |
| Alaska |  |  |  |  |  |  |  |  |  |  |  |  |
| Arizona |  |  |  |  |  |  |  |  |  |  |  |  |
| Arkansas |  |  |  |  |  |  |  |  |  | 2 | 1 |  |
| California |  |  |  |  |  |  |  |  |  |  |  |  |
| Colorado |  |  |  |  |  |  |  |  |  |  |  |  |
| Connecticut |  |  |  |  |  |  |  |  |  |  |  |  |
| Delaware |  |  |  |  |  |  |  |  |  |  |  |  |
| Florida |  |  |  |  |  |  |  |  |  |  |  |  |
| Georgia |  |  |  |  |  |  |  |  |  |  |  |  |
| Hawaii |  |  |  |  |  |  |  |  |  |  |  |  |
| Idaho |  |  |  |  |  |  |  |  |  |  |  |  |
| Illinois |  |  |  |  |  |  |  |  |  |  |  |  |
| Indiana |  |  |  |  |  |  |  |  |  |  |  |  |
| Iowa |  |  |  |  |  |  |  |  |  |  |  |  |
| Kansas |  |  |  |  |  |  |  |  |  |  |  |  |
| Kentucky |  |  |  |  |  |  |  |  |  |  |  |  |
| Louisiana |  |  |  |  |  |  |  |  |  |  |  |  |
| Maine |  |  |  |  |  |  |  |  |  |  |  |  |
| Maryland |  |  |  |  |  |  |  |  |  |  |  |  |
| Massachusetts |  |  | 1 |  |  |  |  |  |  |  | 1 |  |
| Michigan |  |  |  |  |  |  |  |  |  |  |  |  |
| Minnesota |  |  |  |  |  | 1 |  |  |  |  | 1 |  |
| Mississippi |  |  |  |  |  |  |  |  |  |  |  |  |
| Missouri |  |  |  |  |  |  |  |  |  |  |  |  |
| Montana |  |  |  |  |  |  |  |  |  |  |  |  |
| Nebraska |  |  |  |  |  |  |  |  |  |  |  |  |
| Nevada |  |  |  |  |  |  |  |  |  |  |  |  |
| New Hampshire |  |  |  |  |  |  |  |  |  |  |  |  |
| New Jersey |  |  |  |  |  |  |  |  |  |  |  |  |
| New Mexico |  |  |  |  |  |  |  |  |  |  |  |  |
| New York |  |  |  |  |  |  |  |  |  |  |  | 2 |
| North Carolina |  |  |  |  |  |  |  |  |  |  |  |  |
| North Dakota |  |  |  |  |  |  |  |  |  |  |  |  |
| Ohio |  |  |  |  |  |  |  |  |  |  |  |  |
| Oklahoma |  |  |  |  |  |  |  |  |  |  |  |  |
| Oregon |  |  |  |  |  |  |  |  |  |  |  |  |
| Pennsylvania |  |  |  |  |  |  |  |  |  |  |  |  |
| Rhode Island |  |  |  |  |  |  |  |  |  |  |  |  |
| South Carolina |  |  |  |  |  |  |  |  |  |  |  |  |
| South Dakota |  |  |  |  |  |  |  |  |  |  |  |  |
| Tennessee |  |  |  |  |  |  |  |  |  |  |  |  |
| Texas |  | 2 |  |  |  |  |  | 1 |  |  |  |  |
| Utah |  |  |  |  |  |  |  |  |  |  |  |  |
| Vermont |  |  |  |  |  |  |  |  |  |  |  |  |
| Virginia |  |  |  |  |  |  |  |  |  |  |  |  |
| Washington |  |  |  |  |  |  |  |  |  |  |  |  |
| West Virginia |  |  |  |  |  |  |  |  |  |  |  |  |
| Wisconsin |  |  |  |  |  |  |  |  |  |  |  |  |
| Wyoming |  |  |  |  |  |  |  |  |  |  |  |  |
| **State** | **2005 H1** | **2005 H2** | **2006 H1** | **2006 H2** | **2007 H1** | **2007 H2** | **2008 H1** | **2008 H2** | **2009 H1** | **2009 H2** | **2010 H1** | **2010 H2** |
| Alabama |  |  |  |  |  |  |  |  |  |  |  |  |
| Alaska |  |  |  |  |  |  |  |  |  |  |  |  |
| Arizona |  |  |  | 1 |  |  |  |  |  |  |  |  |
| Arkansas | 1 |  |  |  | 2 |  |  |  |  |  |  |  |
| California |  |  | 1 |  |  |  |  |  |  |  |  |  |
| Colorado |  |  |  |  |  |  |  |  |  |  |  |  |
| Connecticut |  | 1 | 7 | 7 | 1 |  | 11 | 2 |  | 3 | 5 | 2 |
| Delaware |  |  |  |  | 1 |  |  |  |  |  |  |  |
| Florida | 1 |  |  | 1 |  | 1 | 3 | 2 | 2 | 1 | 6 |  |
| Georgia |  |  |  |  |  |  |  |  |  |  |  |  |
| Hawaii |  |  |  |  |  |  |  |  |  |  |  |  |
| Idaho |  |  |  |  |  |  |  |  |  |  |  |  |
| Illinois |  |  |  |  |  |  |  |  |  |  |  |  |
| Indiana |  |  | 1 |  |  |  |  |  |  |  |  |  |
| Iowa |  |  |  |  |  |  |  |  |  |  |  |  |
| Kansas |  |  |  |  |  |  |  |  |  |  |  |  |
| Kentucky |  |  |  |  |  |  |  |  | 1 |  |  |  |
| Louisiana |  |  |  | 1 |  |  |  |  |  |  |  |  |
| Maine |  |  |  |  |  | 1 |  |  |  |  |  |  |
| Maryland |  |  |  |  |  |  |  |  |  |  |  |  |
| Massachusetts | 1 | 1 | 6 | 7 | 1 | 1 |  |  |  |  |  | 1 |
| Michigan |  |  |  |  |  |  |  |  |  | 1 |  |  |
| Minnesota |  |  |  |  |  |  |  |  |  |  |  |  |
| Mississippi |  |  |  |  |  |  |  |  |  |  |  |  |
| Missouri |  |  |  |  |  |  | 5 |  |  | 1 |  | 1 |
| Montana |  |  |  |  |  |  |  |  |  |  |  |  |
| Nebraska |  |  |  |  |  |  |  |  |  |  |  |  |
| Nevada |  |  |  |  |  |  |  |  |  |  |  |  |
| New Hampshire |  |  |  |  |  |  |  | 1 |  |  |  |  |
| New Jersey |  | 1 |  | 1 |  | 1 |  | 1 |  |  | 2 |  |
| New Mexico |  |  |  |  |  |  |  |  |  |  |  |  |
| New York |  |  | 3 |  | 1 | 1 | 5 |  |  | 2 | 8 | 1 |
| North Carolina |  |  |  |  |  |  |  |  |  |  |  |  |
| North Dakota |  |  |  |  |  |  |  |  |  |  |  |  |
| Ohio |  |  |  |  |  |  |  |  | 10 | 1 |  |  |
| Oklahoma |  |  |  |  |  |  |  |  |  |  |  |  |
| Oregon |  |  |  |  |  |  |  |  |  |  |  |  |
| Pennsylvania |  |  |  |  | 2 |  | 3 | 1 | 1 |  |  | 3 |
| Rhode Island |  |  | 1 |  |  |  |  |  |  |  |  |  |
| South Carolina |  |  |  |  |  |  |  |  |  |  |  |  |
| South Dakota |  |  |  |  |  |  |  |  |  |  |  |  |
| Tennessee |  |  |  |  |  |  |  |  |  |  |  |  |
| Texas |  |  |  | 1 |  | 1 | 2 |  |  |  |  |  |
| Utah |  |  |  |  |  |  |  |  |  |  |  |  |
| Vermont |  |  |  |  |  |  |  | 1 |  |  |  |  |
| Virginia |  |  | 1 |  |  |  |  |  |  | 2 |  |  |
| Washington |  |  |  |  |  |  |  |  |  |  |  |  |
| West Virginia |  |  |  |  |  |  |  |  |  |  |  |  |
| Wisconsin |  | 2 |  |  | 1 |  |  |  |  |  |  |  |
| Wyoming |  |  |  |  |  |  |  |  |  |  |  |  |
| **State** | **2011 H1** | **2011 H2** | **2012 H1** | **2012 H2** | **2013 H1** | **2013 H2** | **2014 H1** | **2014 H2** | **2015 H1** | **2015 H2** | **2016 H1** | **2016 H2** |
| Alabama |  |  |  |  |  |  |  |  |  |  |  | 1 |
| Alaska |  |  |  |  |  |  |  |  |  |  |  |  |
| Arizona |  |  |  |  |  |  |  |  |  |  |  |  |
| Arkansas |  |  |  |  |  |  | 3 | 5 | 3 | 2 | 1 |  |
| California |  |  |  |  |  |  |  |  |  | 1 | 2 |  |
| Colorado |  |  |  |  |  |  |  |  |  |  |  |  |
| Connecticut | 4 | 1 |  | 1 |  | 1 |  | 13 | 3 | 5 | 4 | 2 |
| Delaware |  |  |  |  | 1 | 1 |  |  |  |  |  | 2 |
| Florida | 5 |  | 2 | 3 | 1 |  | 2 | 8 | 1 | 10 | 5 | 54 |
| Georgia |  |  |  |  |  |  |  |  |  |  |  |  |
| Hawaii |  |  |  |  |  |  |  |  |  |  |  |  |
| Idaho |  |  |  |  |  |  |  |  |  |  |  |  |
| Illinois |  |  |  |  |  |  |  |  | 1 |  |  |  |
| Indiana |  |  |  |  |  |  |  |  |  |  |  |  |
| Iowa |  |  |  |  |  | 1 |  |  |  |  |  |  |
| Kansas |  |  |  |  |  |  |  |  |  |  |  |  |
| Kentucky |  |  |  |  |  |  | 1 | 2 | 2 | 3 |  | 2 |
| Louisiana |  |  |  |  |  | 7 | 10 | 4 | 7 | 8 | 6 | 7 |
| Maine |  |  |  |  |  |  | 1 |  |  |  |  |  |
| Maryland |  |  |  |  | 1 |  | 2 |  |  | 1 | 2 |  |
| Massachusetts |  |  | 1 | 3 | 5 | 3 | 4 | 2 |  | 2 | 10 | 8 |
| Michigan |  |  |  |  |  |  |  |  |  |  |  |  |
| Minnesota |  |  |  |  |  |  |  |  |  |  |  |  |
| Mississippi |  |  |  |  |  |  |  |  |  |  |  |  |
| Missouri |  |  |  |  |  |  |  |  |  |  |  |  |
| Montana |  |  |  |  |  |  |  |  |  |  |  |  |
| Nebraska |  |  |  |  |  |  |  |  |  |  |  |  |
| Nevada |  |  |  |  |  |  |  |  |  |  |  | 1 |
| New Hampshire |  |  |  |  |  | 1 |  |  |  |  |  |  |
| New Jersey | 4 | 2 |  |  |  | 3 | 1 | 1 | 3 | 10 | 33 | 33 |
| New Mexico |  |  |  |  | 1 |  |  |  |  |  |  |  |
| New York | 6 | 2 |  |  | 1 | 2 | 7 | 5 | 3 | 7 | 8 | 8 |
| North Carolina |  |  |  |  |  |  |  |  |  |  | 1 |  |
| North Dakota |  |  |  |  |  |  |  |  |  |  |  |  |
| Ohio |  |  |  |  |  |  |  |  |  |  |  |  |
| Oklahoma |  |  |  |  |  |  |  |  |  | 1 |  |  |
| Oregon |  |  |  |  |  |  |  |  |  |  |  |  |
| Pennsylvania | 4 | 7 | 3 | 1 | 3 |  | 2 |  | 4 | 3 | 15 | 15 |
| Rhode Island |  |  |  |  |  |  |  |  |  | 1 | 5 |  |
| South Carolina |  |  |  |  |  |  |  |  |  | 2 | 3 |  |
| South Dakota |  |  |  |  |  |  |  |  |  |  |  |  |
| Tennessee |  |  |  |  |  |  |  | 2 | 4 | 4 | 4 | 1 |
| Texas |  |  | 1 |  |  |  |  |  |  |  | 2 |  |
| Utah |  |  |  |  |  |  |  |  |  |  |  |  |
| Vermont |  |  |  |  |  | 1 |  |  |  |  |  |  |
| Virginia |  |  | 1 |  | 1 |  |  | 3 | 2 |  |  |  |
| Washington |  |  |  |  |  |  |  |  |  |  |  |  |
| West Virginia |  |  |  | 2 |  |  | 3 |  | 3 |  |  |  |
| Wisconsin |  |  |  |  | 1 |  |  |  |  |  |  |  |
| Wyoming |  |  |  |  |  |  |  |  |  |  |  |  |
| **State** | **2017 H1** | **2017 H2** | **2018 H1** | **2018 H2** | **2019 H1** | **2019 H2** | **2020 H1** | **2020 H2** | **2021 H1** | **2021 H2** | **2022 H1** | **2022 H2** |
| Alabama |  |  |  | 1 | 3 | 2 |  | 2 | 3 | 12 | 20 | 22 |
| Alaska |  |  |  |  |  |  | 1 |  |  | 6 | 4 | 8 |
| Arizona | 1 |  | 2 |  | 2 | 5 | 1 | 2 | 4 | 13 | 25 | 3 |
| Arkansas | 1 | 6 |  | 1 |  | 2 | 1 | 4 | 6 | 16 | 35 | 14 |
| California | 6 | 8 | 9 | 8 | 12 | 28 | 24 | 61 | 85 | 69 | 74 | 59 |
| Colorado |  |  | 1 |  |  |  |  |  | 1 | 4 | 8 | 12 |
| Connecticut | 22 | 4 | 11 | 7 | 16 | 44 | 33 | 24 | 54 | 83 | 116 | 157 |
| Delaware | 1 | 4 | 3 | 5 | 13 | 5 | 1 | 5 | 13 | 12 | 30 | 23 |
| Florida | 76 | 95 | 121 | 97 | 83 | 159 | 185 | 191 | 251 | 295 | 459 | 411 |
| Georgia |  |  |  |  | 1 | 1 | 2 | 2 | 22 | 51 | 34 | 22 |
| Hawaii |  |  |  | 1 |  |  |  | 1 |  | 2 | 3 |  |
| Idaho |  |  |  |  |  |  |  |  |  | 1 | 3 | 3 |
| Illinois |  | 1 |  |  | 3 | 8 | 7 | 19 | 18 | 9 | 25 | 41 |
| Indiana |  | 6 | 3 | 1 | 17 | 14 | 6 | 25 | 63 | 59 | 152 | 137 |
| Iowa |  |  | 2 |  |  | 1 |  |  |  |  | 1 |  |
| Kansas |  |  |  |  |  | 2 | 1 |  | 1 | 5 | 108 | 15 |
| Kentucky |  |  | 1 | 2 | 2 | 11 | 24 | 25 | 15 | 47 | 28 | 40 |
| Louisiana | 10 | 6 |  | 2 | 1 | 9 | 7 | 4 | 2 | 11 | 8 | 7 |
| Maine |  |  |  |  |  | 3 | 9 | 1 | 5 | 6 | 1 | 2 |
| Maryland | 5 |  | 2 | 29 | 2 | 10 | 10 | 39 | 752 | 799 | 690 | 488 |
| Massachusetts | 8 | 4 | 3 | 3 | 12 | 20 | 15 | 34 | 41 | 48 | 80 | 123 |
| Michigan |  |  |  |  | 2 | 2 | 3 | 18 | 9 | 16 | 25 | 30 |
| Minnesota |  |  |  |  |  | 2 | 6 |  | 7 | 7 | 42 | 141 |
| Mississippi |  | 2 |  |  |  |  |  | 3 | 5 | 3 | 9 | 4 |
| Missouri | 4 |  | 6 | 5 | 6 | 9 | 11 | 15 | 16 | 16 | 21 | 47 |
| Montana |  |  |  |  |  |  |  |  | 4 | 7 | 12 | 13 |
| Nebraska |  |  |  |  |  |  |  | 1 | 2 |  | 7 | 3 |
| Nevada |  |  |  |  |  | 2 | 2 | 2 | 3 | 3 | 14 | 4 |
| New Hampshire |  |  |  |  | 10 | 27 | 20 | 17 | 68 | 48 | 74 | 108 |
| New Jersey | 31 | 14 | 33 | 52 | 187 | 275 | 448 | 1,042 | 1,228 | 1,373 | 1,391 | 1,458 |
| New Mexico | 2 |  |  |  |  |  |  | 2 | 8 | 4 | 17 | 4 |
| New York | 13 | 23 | 36 | 8 | 24 | 46 | 62 | 58 | 147 | 109 | 199 | 159 |
| North Carolina | 1 | 1 | 6 | 3 |  | 1 | 6 | 5 | 8 | 71 | 98 | 157 |
| North Dakota |  |  |  |  |  | 3 |  |  | 6 | 6 | 7 | 11 |
| Ohio |  |  | 6 | 14 | 40 | 133 | 165 | 362 | 669 | 648 | 600 | 688 |
| Oklahoma |  |  | 1 |  |  |  | 1 |  | 1 | 4 | 8 | 18 |
| Oregon |  |  |  |  |  |  |  |  | 1 | 14 | 30 | 9 |
| Pennsylvania | 15 | 20 | 21 | 24 | 86 | 209 | 87 | 90 | 161 | 185 | 251 | 190 |
| Rhode Island |  | 1 |  |  | 4 | 1 |  | 27 | 126 | 113 | 134 | 122 |
| South Carolina | 2 |  | 2 | 1 |  |  | 3 | 6 | 3 | 9 | 64 | 75 |
| South Dakota |  |  |  |  |  |  |  |  |  |  |  |  |
| Tennessee |  |  |  | 2 | 1 | 11 | 10 | 15 | 20 | 57 | 224 | 317 |
| Texas | 1 |  | 1 | 1 | 9 | 1 | 16 | 15 | 23 | 29 | 53 | 75 |
| Utah |  |  |  |  |  |  |  |  |  | 1 | 5 | 2 |
| Vermont |  |  | 5 |  |  |  | 2 | 12 | 4 | 7 | 22 | 23 |
| Virginia | 1 | 2 | 2 | 4 | 1 | 4 | 8 | 58 | 324 | 618 | 726 | 616 |
| Washington |  |  |  |  |  |  | 11 | 2 | 14 | 28 | 45 | 51 |
| West Virginia | 2 | 3 | 10 | 3 | 7 | 10 | 6 | 14 | 85 | 57 | 56 | 99 |
| Wisconsin |  |  | 27 | 26 | 27 | 20 | 22 | 8 | 11 | 19 | 32 | 12 |
| Wyoming |  |  |  |  |  |  |  |  |  |  |  |  |
| **State** | **2023 H1** | **2023 H2** | **2024 H1** | **2024 H2** | **2025 H1** | **2025 H2** |  |  |  |  |  |  |
| Alabama | 24 | 25 | 25 | 29 | 30 | 28 |  |  |  |  |  |  |
| Alaska | 5 | 3 | 33 | 39 | 19 | 14 |  |  |  |  |  |  |
| Arizona | 9 | 9 | 44 | 275 | 178 | 160 |  |  |  |  |  |  |
| Arkansas | 18 | 35 | 63 | 122 | 59 | 23 |  |  |  |  |  |  |
| California | 81 | 62 | 166 | 175 | 141 | 100 |  |  |  |  |  |  |
| Colorado | 8 | 3 | 5 | 6 | 14 | 4 |  |  |  |  |  |  |
| Connecticut | 190 | 223 | 222 | 307 | 172 | 257 |  |  |  |  |  |  |
| Delaware | 44 | 23 | 20 | 10 | 2 | 1 |  |  |  |  |  |  |
| Florida | 481 | 390 | 372 | 489 | 384 | 190 |  |  |  |  |  |  |
| Georgia | 65 | 47 | 42 | 37 | 48 | 59 |  |  |  |  |  |  |
| Hawaii | 1 | 5 | 1 |  | 5 | 3 |  |  |  |  |  |  |
| Idaho |  | 13 | 42 | 40 | 11 | 7 |  |  |  |  |  |  |
| Illinois | 20 | 49 | 51 | 40 | 59 | 26 |  |  |  |  |  |  |
| Indiana | 181 | 186 | 177 | 294 | 328 | 96 |  |  |  |  |  |  |
| Iowa | 33 | 58 | 212 | 133 | 118 | 55 |  |  |  |  |  |  |
| Kansas | 16 | 7 | 17 | 12 | 8 | 1 |  |  |  |  |  |  |
| Kentucky | 47 | 50 | 33 | 77 | 40 | 23 |  |  |  |  |  |  |
| Louisiana | 65 | 28 | 57 | 31 | 28 | 45 |  |  |  |  |  |  |
| Maine | 10 | 9 | 17 | 13 | 17 | 11 |  |  |  |  |  |  |
| Maryland | 443 | 442 | 680 | 470 | 266 | 92 |  |  |  |  |  |  |
| Massachusetts | 184 | 228 | 248 | 246 | 116 | 164 |  |  |  |  |  |  |
| Michigan | 48 | 41 | 47 | 34 | 46 | 35 |  |  |  |  |  |  |
| Minnesota | 87 | 139 | 93 | 61 | 20 | 31 |  |  |  |  |  |  |
| Mississippi | 2 | 2 | 25 | 16 | 8 | 5 |  |  |  |  |  |  |
| Missouri | 107 | 159 | 381 | 432 | 228 | 108 |  |  |  |  |  |  |
| Montana | 23 | 29 | 57 | 110 | 83 | 35 |  |  |  |  |  |  |
| Nebraska | 11 | 5 | 4 | 2 | 3 | 2 |  |  |  |  |  |  |
| Nevada | 5 | 1 | 12 | 7 | 2 | 2 |  |  |  |  |  |  |
| New Hampshire | 109 | 93 | 149 | 216 | 86 | 37 |  |  |  |  |  |  |
| New Jersey | 1,878 | 2,115 | 1,918 | 293 | 284 | 100 |  |  |  |  |  |  |
| New Mexico | 13 | 14 | 12 | 12 | 7 | 10 |  |  |  |  |  |  |
| New York | 151 | 112 | 203 | 149 | 166 | 61 |  |  |  |  |  |  |
| North Carolina | 238 | 339 | 538 | 802 | 535 | 251 |  |  |  |  |  |  |
| North Dakota | 19 | 11 | 25 | 57 | 22 | 84 |  |  |  |  |  |  |
| Ohio | 2,187 | 2,978 | 3,416 | 3,164 | 2,562 | 2,187 |  |  |  |  |  |  |
| Oklahoma | 10 | 21 | 113 | 198 | 69 | 82 |  |  |  |  |  |  |
| Oregon | 9 | 14 | 36 | 82 | 37 | 16 |  |  |  |  |  |  |
| Pennsylvania | 2,191 | 3,459 | 5,077 | 2,900 | 1,999 | 1,098 |  |  |  |  |  |  |
| Rhode Island | 179 | 145 | 187 | 132 | 139 | 122 |  |  |  |  |  |  |
| South Carolina | 81 | 30 | 60 | 221 | 634 | 310 |  |  |  |  |  |  |
| South Dakota |  |  |  | 5 |  | 2 |  |  |  |  |  |  |
| Tennessee | 350 | 435 | 585 | 667 | 514 | 170 |  |  |  |  |  |  |
| Texas | 57 | 48 | 52 | 62 | 59 | 41 |  |  |  |  |  |  |
| Utah |  |  | 4 | 2 |  |  |  |  |  |  |  |  |
| Vermont | 31 | 27 | 26 | 43 | 3 | 2 |  |  |  |  |  |  |
| Virginia | 658 | 590 | 768 | 926 | 813 | 400 |  |  |  |  |  |  |
| Washington | 42 | 115 | 176 | 213 | 132 | 93 |  |  |  |  |  |  |
| West Virginia | 378 | 516 | 644 | 686 | 502 | 262 |  |  |  |  |  |  |
| Wisconsin | 45 | 69 | 102 | 85 | 27 | 18 |  |  |  |  |  |  |
| Wyoming |  | 1 |  | 10 |  |  |  |  |  |  |  |  |

**Caption.** State-semiannual period level drug reports from the National Forensic Laboratory Information System (NFLIS). H1 refers to January through June, and H2 refers to July through December. NFLIS data were obtained on May 7, 2026.

**eTable 2. Number of Medetomidine Reports in NFLIS by Semiannual Period, 1999–2025**

| **State** | **Semiannual Period** | | | | | | | | |
| --- | --- | --- | --- | --- | --- | --- | --- | --- | --- |
|  | **2021 H2** | **2022 H1** | **2022 H2** | **2023 H1** | **2023 H2** | **2024 H1** | **2024 H1** | **2025 H1** | **2025 H2** |
| Alabama |  |  |  |  |  |  |  | 2 | 5 |
| Alaska |  |  |  |  |  |  |  |  | 3 |
| Arizona |  |  |  |  |  |  |  |  | 1 |
| Arkansas |  |  |  |  |  | 2 | 2 | 4 | 4 |
| California |  |  |  |  |  |  |  | 4 | 13 |
| Colorado |  |  |  |  |  |  |  | 1 | 4 |
| Connecticut |  |  |  |  |  |  | 21 | 76 | 127 |
| Delaware |  |  | 1 |  |  |  | 2 | 6 | 7 |
| Florida |  |  |  | 1 |  |  | 5 | 42 | 19 |
| Georgia |  |  |  |  |  |  | 1 | 4 | 7 |
| Illinois |  |  |  |  |  |  | 9 | 24 | 14 |
| Indiana |  |  |  | 2 |  | 5 | 89 | 264 | 174 |
| Iowa |  |  |  |  |  | 3 | 20 | 76 | 43 |
| Kansas |  |  |  |  |  |  | 1 | 11 | 2 |
| Kentucky |  |  |  |  |  |  | 9 | 29 | 21 |
| Louisiana |  |  |  |  |  |  |  | 2 | 2 |
| Maine |  |  |  |  |  |  |  | 5 | 5 |
| Maryland |  | 2 | 22 | 5 | 7 | 6 | 3 | 53 | 67 |
| Massachusetts |  |  |  |  |  |  | 3 | 25 | 14 |
| Michigan |  |  |  |  |  |  | 10 | 23 | 15 |
| Minnesota |  |  |  |  |  |  | 1 | 22 | 39 |
| Mississippi |  |  |  |  |  |  |  |  | 5 |
| Missouri |  |  |  |  |  |  | 17 | 71 | 48 |
| Montana |  |  |  |  |  |  |  |  | 2 |
| Nebraska |  |  |  |  |  |  |  |  | 1 |
| Nevada |  |  |  |  |  |  |  |  | 2 |
| New Hampshire |  |  |  |  |  |  | 3 | 11 | 12 |
| New Jersey |  |  |  | 3 |  | 6 | 145 | 205 | 72 |
| New Mexico |  |  |  |  |  |  |  |  | 2 |
| New York |  |  |  |  |  |  | 9 | 64 | 37 |
| North Carolina |  |  |  |  |  |  | 32 | 136 | 114 |
| North Dakota |  |  |  |  | 1 |  | 2 | 6 | 23 |
| Ohio | 10 | 9 | 158 | 103 | 86 | 52 | 318 | 935 | 929 |
| Oklahoma |  |  |  |  |  |  |  | 12 | 14 |
| Oregon |  |  |  |  |  |  |  | 1 | 2 |
| Pennsylvania |  |  |  | 1 | 2 | 71 | 1,638 | 2,303 | 1,662 |
| Rhode Island |  |  |  |  |  |  |  | 37 | 58 |
| South Carolina |  |  |  |  |  | 3 | 4 | 19 | 45 |
| South Dakota |  |  |  |  |  |  |  |  | 1 |
| Tennessee |  |  |  |  |  |  | 22 | 79 | 111 |
| Texas |  |  |  |  |  |  |  | 20 | 13 |
| Vermont |  |  |  |  |  |  |  |  | 2 |
| Virginia | 2 | 28 | 43 | 12 | 14 | 10 | 75 | 262 | 221 |
| Washington |  |  |  |  |  |  |  | 3 | 32 |
| West Virginia |  |  |  |  | 4 | 3 | 17 | 47 | 47 |
| Wisconsin |  |  |  |  |  |  |  | 15 | 9 |
| Wyoming |  |  |  |  | 1 |  |  |  |  |

**Caption.** State-semiannual period level drug reports from the National Forensic Laboratory Information System (NFLIS). H1 refers to January through June, and H2 refers to July through December. NFLIS data were obtained on May 7, 2026.

**eTable 3. Changes in Mean Xylazine and Medetomidine Report Rates Relative to Index Semiannual Period of Xylazine Scheduling**

| **Semiannual Period Relative to Xylazine Scheduling** | **Mean xylazine reports per 100,000 NFLIS reports (95% CI)** | | **Mean medetomidine reports per 100,000 NFLIS reports (95% CI)** | |
| --- | --- | --- | --- | --- |
|  | **Treatment group** | **Control group** | **Treatment group** | **Control group** |
| -5 | 940.57  (688.56, 1438.01) | 778.99  (607.92, 1155.92) | 0.00  (0.00, 0.00) | 1.59  (1.09, 4.46) |
| -4 | 2903.48  (2127.34, 4772.79) | 1183.66  (923.65, 1741.57) | 0.00  (0.00, 0.00) | 0.63  (0.43, 1.64) |
| -3 | 3262.50  (2367.59, 5372.72) | 1429.33  (1124.96, 2054.14) | 2.64  (1.69, 5.93) | 4.10  (2.87, 10.67) |
| -2 | 4290.07  (3099.57, 7281.93) | 1802.71  (1454.27, 2430.02) | 2.64  (1.70, 5.95) | 2.15  (1.51, 4.99) |
| -1 | 4335.91  (3089.42, 7334.15) | 1988.06  (1592.68, 2617.23) | 160.65  (111.22, 296.40) | 29.23  (22.09, 45.70) |
| 0 | 7131.69  (5351.80, 10113.09) | 2337.18  (1879.20, 3041.75) | 37.57  (26.01, 75.97) | 74.83  (59.75, 100.71) |
| +1 | 10824.96  (8452.12, 14050.98) | 2581.51  (2109.84, 3354.75) | 56.38  (39.03, 92.69) | 153.19  (124.72, 210.10) |
| +2 | 12563.91  (9641.12, 15479.88) | 3105.69  (2538.48, 4046.23) | 100.96  (67.40, 149.70) | 164.82  (131.82, 220.49) |
| +3 | 10793.57  (8780.48, 12216.83) | 3015.72  (2507.21, 3908.24) | 2112.25  (1532.39, 3339.19) | 356.27  (287.48, 493.64) |
| +4 | 8172.82  (6254.25, 9815.28) | 2113.07  (1770.72, 2591.59) | 4796.82  (3470.44, 6666.32) | 534.81  (428.20, 719.32) |
| +5 | 6601.55  (4978.87, 8151.58) | 1800.67  (1494.66, 2415.57) | 5783.23  (4213.42, 7686.22) | 823.21  (670.51, 1120.50) |

**Caption.** Mean xylazine and medetomidine reports per 100,000 state-level drug reports in the National Forensic Laboratory Information System (NFLIS). The treatment group includes states that scheduled xylazine, while the control group includes states that did not. NFLIS data were obtained on May 7, 2026.

**eTable 4. Difference-in-Differences and Event-Study Coefficients of Changes in Xylazine and Medetomidine Report Rates Following State Xylazine Scheduling**

| **Model** | **Semiannual Period Relative to Xylazine Scheduling** | **Estimate (95% CI)** | ***P Value*** |
| --- | --- | --- | --- |
| Primary analysis, xylazine | **ATT** | 2872.29 (-2024.63, 7769.21) | .250 |
|  | **Event study coefficients** |  |  |
|  | T_–5_ | 668.49 (-110.79, 1447.77) | .093 |
|  | T_–4_ | 1503.00 (-293.07, 3299.07) | .101 |
|  | T_–3_ | 94.76 (-320.24, 509.77) | .654 |
|  | T_–2_ | 958.30 (-715.47, 2632.07) | .262 |
|  | T_–1_ | -36.13 (-543.57, 471.31) | .889 |
|  | T_0_ | 2783.16 (543.75, 5022.57) | .015 |
|  | T_1_ | 6194.35 (1505.69, 10883.01) | .010 |
|  | T_2_ | 5794.91 (-2718.11, 14307.94) | .182 |
|  | T_3_ | 2462.58 (-6669.59, 11594.75) | .597 |
|  | T_4_ | 1128.61 (-7027.93, 9285.15) | .786 |
|  | T_5_ | -118.63 (-7836.04, 7598.78) | .976 |
| Primary analysis, medetomidine | **ATT** | 1536.51 (211.14, 2861.88) | .023 |
|  | **Event study coefficients** |  |  |
|  | T_–4_ | -0.11 (-0.40, 0.18) | .475 |
|  | T_–3_ | 3.01 (-3.35, 9.37) | .354 |
|  | T_–2_ | -1.07 (-3.79, 1.65) | .441 |
|  | T_–1_ | 185.60 (-59.84, 431.04) | .138 |
|  | T_0_ | -144.04 (-381.29, 93.22) | .234 |
|  | T_1_ | -139.19 (-387.76, 109.39) | .272 |
|  | T_2_ | -160.60 (-544.34, 223.14) | .412 |
|  | T_3_ | 2024.27 (-812.41, 4860.94) | .162 |
|  | T_4_ | 4839.59 (1273.79, 8405.40) | .008 |
|  | T_5_ | 5677.21 (1979.81, 9374.61) | .003 |
| Sensitivity analysis (excluding Florida), medetomidine | **ATT** | 3668.18 (-3175.77, 10512.14) | .293 |
|  | **Event study coefficients** |  |  |
|  | T_–5_ | 761.89 (-107.25, 1631.02) | .086 |
|  | T_–4_ | 1710.30 (-296.58, 3717.19) | .095 |
|  | T_–3_ | 116.71 (-355.02, 588.44) | .628 |
|  | T_–2_ | 1085.64 (-808.81, 2980.09) | .261 |
|  | T_–1_ | -35.77 (-615.72, 544.17) | .904 |
|  | T_0_ | 3130.49 (676.03, 5584.95) | .012 |
|  | T_1_ | 7021.79 (1946.77, 12096.82) | .007 |
|  | T_2_ | 6839.81 (-3126.38, 16805.99) | .179 |
|  | T_3_ | 2844.79 (-8083.00, 13772.59) | .610 |
|  | T_4_ | 1261.85 (-8521.76, 11045.45) | .800 |
|  | T_5_ | -215.99 (-9474.02, 9042.04) | .964 |
| Sensitivity analysis (excluding Florida), medetomidine | **ATT** | 1762.35 (333.49, 3191.20) | .016 |
|  | **Event study coefficients** |  |  |
|  | T_–4_ | -0.11 (-0.40, 0.18) | .475 |
|  | T_–3_ | 3.01 (-3.35, 9.37) | .354 |
|  | T_–2_ | -1.07 (-3.79, 1.65) | .441 |
|  | T_–1_ | 185.60 (-59.84, 431.04) | .138 |
|  | T_0_ | -144.04 (-381.29, 93.22) | .234 |
|  | T_1_ | -139.19 (-387.76, 109.39) | .272 |
|  | T_2_ | -160.60 (-544.34, 223.14) | .412 |
|  | T_3_ | 2024.27 (-812.41, 4860.94) | .162 |
|  | T_4_ | 4839.59 (1273.79, 8405.40) | .008 |
|  | T_5_ | 5677.21 (1979.81, 9374.61) | .003 |
| Sensitivity analysis (expanded treatment definition), xylazine | **ATT** | 2359.09 (-1793.17, 6511.35) | .265 |
|  | **Event study coefficients** |  |  |
|  | T_–5_ | 373.11 (-202.95, 949.18) | .204 |
|  | T_–4_ | 902.97 (-220.01, 2025.96) | .115 |
|  | T_–3_ | -3.12 (-459.05, 452.81) | .989 |
|  | T_–2_ | 661.97 (-408.10, 1732.04) | .225 |
|  | T_–1_ | -3.39 (-405.70, 398.92) | .987 |
|  | T_0_ | 1874.49 (189.82, 3559.15) | .029 |
|  | T_1_ | 4368.44 (937.38, 7799.49) | .013 |
|  | T_2_ | 4402.58 (-2190.16, 10995.33) | .191 |
|  | T_3_ | 1611.83 (-8463.22, 11686.88) | .754 |
|  | T_4_ | 1128.77 (-7090.61, 9348.15) | .788 |
|  | T_5_ | -119.40 (-7900.28, 7661.48) | .976 |
| Sensitivity analysis (expanded treatment definition), medetomidine | **ATT** | 1253.87 (134.89, 2372.86) | .028 |
|  | **Event study coefficients** |  |  |
|  | T_–5_ | -0.36 (-1.86, 1.14) | .635 |
|  | T_–4_ | -3.11 (-15.60, 9.37) | .625 |
|  | T_–3_ | -0.88 (-8.48, 6.73) | .821 |
|  | T_–2_ | -6.96 (-32.56, 18.63) | .594 |
|  | T_–1_ | 108.47 (-48.82, 265.76) | .176 |
|  | T_0_ | -70.44 (-212.49, 71.61) | .331 |
|  | T_1_ | 23.73 (-225.47, 272.93) | .852 |
|  | T_2_ | -55.93 (-303.63, 191.78) | .658 |
|  | T_3_ | 1951.37 (-914.07, 4816.81) | .182 |
|  | T_4_ | 4839.59 (1245.39, 8433.80) | .008 |
|  | T_5_ | 5677.21 (1909.84, 9444.57) | .003 |
| Falsification, steroids | **ATT** | 138.34 (-329.40, 606.09) | .562 |
|  | **Event study coefficients** |  |  |
|  | T_–5_ | -519.82 (-1396.02, 356.37) | .245 |
|  | T_–4_ | 483.25 (-273.75, 1240.24) | .211 |
|  | T_–3_ | -116.53 (-662.19, 429.14) | .676 |
|  | T_–2_ | -438.17 (-1062.34, 186.00) | .169 |
|  | T_–1_ | -143.71 (-640.66, 353.25) | .571 |
|  | T_0_ | 351.63 (-259.48, 962.74) | .259 |
|  | T_1_ | 482.29 (-206.88, 1171.46) | .170 |
|  | T_2_ | 70.80 (-235.54, 377.13) | .651 |
|  | T_3_ | 912.20 (-885.50, 2709.89) | .320 |
|  | T_4_ | -118.37 (-440.66, 203.92) | .472 |
|  | T_5_ | 87.87 (-168.99, 344.74) | .503 |
| Falsification, antidepressants | **ATT** | 157.79 (-4.78, 320.37) | .057 |
|  | **Event study coefficients** |  |  |
|  | T_–5_ | 143.95 (-85.65, 373.56) | .219 |
|  | T_–4_ | -145.01 (-439.70, 149.67) | .335 |
|  | T_–3_ | -118.48 (-331.12, 94.15) | .275 |
|  | T_–2_ | 225.81 (-69.95, 521.56) | .135 |
|  | T_–1_ | -100.29 (-285.01, 84.43) | .287 |
|  | T_0_ | 131.68 (-302.34, 565.69) | .552 |
|  | T_1_ | 241.16 (-42.22, 524.54) | .095 |
|  | T_2_ | 73.55 (-49.72, 196.82) | .242 |
|  | T_3_ | 211.24 (-137.56, 560.05) | .235 |
|  | T_4_ | 4.22 (-123.28, 131.72) | .948 |
|  | T_5_ | 46.91 (-98.08, 191.90) | .526 |

**Caption.** ATT=average treatment effect on the treated. T_−5_ indicates five semiannual periods before xylazine scheduling, T_0_ indicates the semiannual period in which xylazine scheduling took effect, and T_5_ indicates five semiannual periods after xylazine scheduling.

**eTable 5. Wald Tests for Pre-Policy Event-Study Coefficients**

| **Model** | **Pre-policy *P* value** |
| --- | --- |
| Primary analysis, xylazine | .091 |
| Primary analysis, medetomidine | .137 |
| Sensitivity analysis (excluding Florida), medetomidine | .084 |
| Sensitivity analysis (excluding Florida), medetomidine | .137 |
| Sensitivity analysis (expanded treatment definition), xylazine | .081 |
| Sensitivity analysis (expanded treatment definition), medetomidine | .209 |
| Falsification, steroids | .094 |
| Falsification, antidepressants | .947 |

**Caption.** Wald tests evaluated the null hypothesis that all pre-policy event-study coefficients are jointly equal to zero. *P*<.05 indicates evidence of differential pre-policy trends before state xylazine scheduling. *P*>.05 supports the parallel trends assumption.

**eFigure 1. State-Level Trends in Xylazine and Medetomidine Report Rates Among States Formally Scheduling Xylazine**

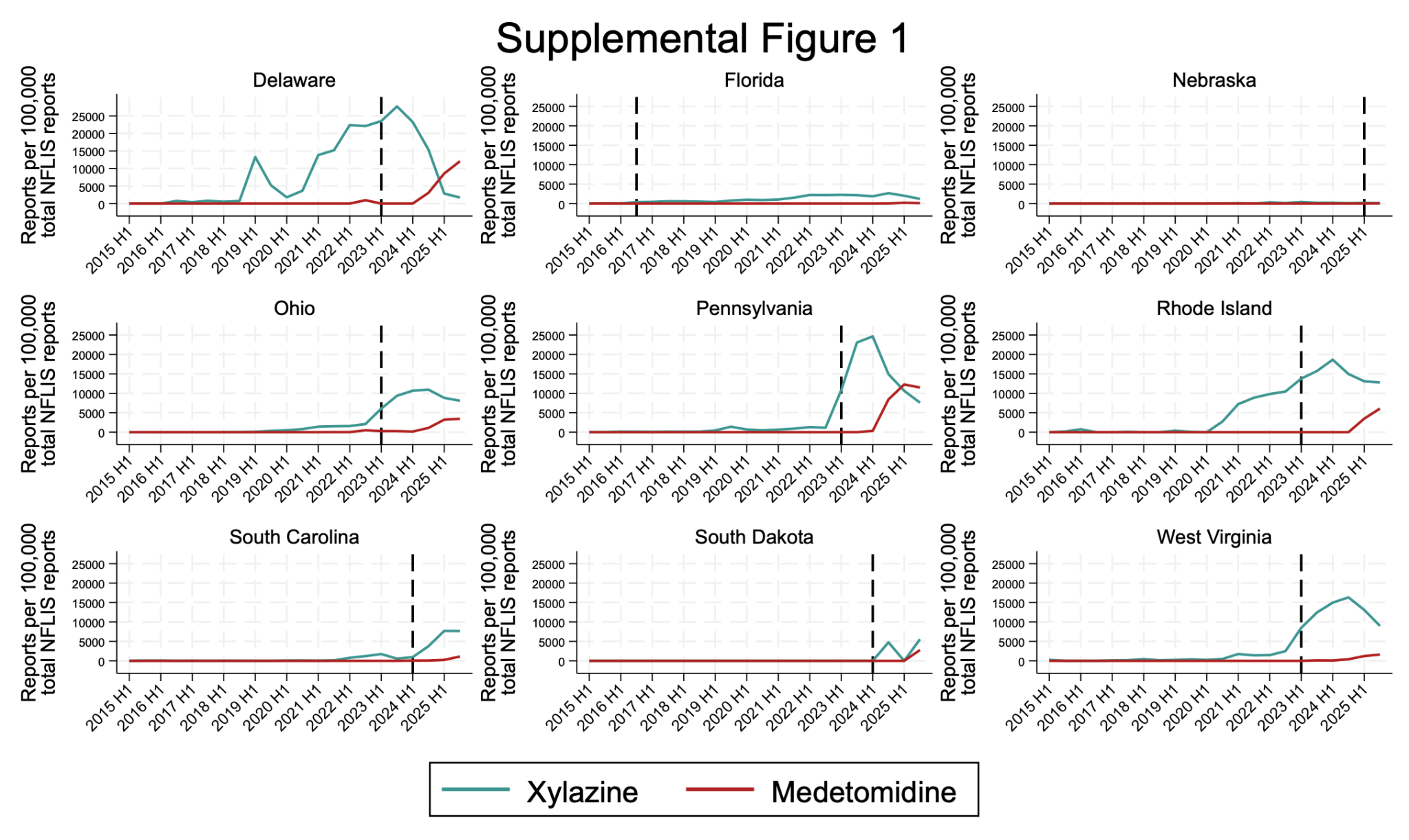

**Caption.** State-level trends in xylazine and medetomidine reports per 100,000 total drug reports in the National Forensic Laboratory Information System (NFLIS) among states that formally scheduled xylazine as a controlled substance during the study period. The teal line represents xylazine reports and the red line represents medetomidine reports. Vertical dashed lines indicate the semiannual period in which xylazine scheduling took effect in each state. NFLIS data were obtained on May 7, 2026.

**eFigure 2. State-Level Trends in Xylazine and Medetomidine Report Rates Among States Enacting Xylazine Criminalization Policies Without Formal Scheduling**

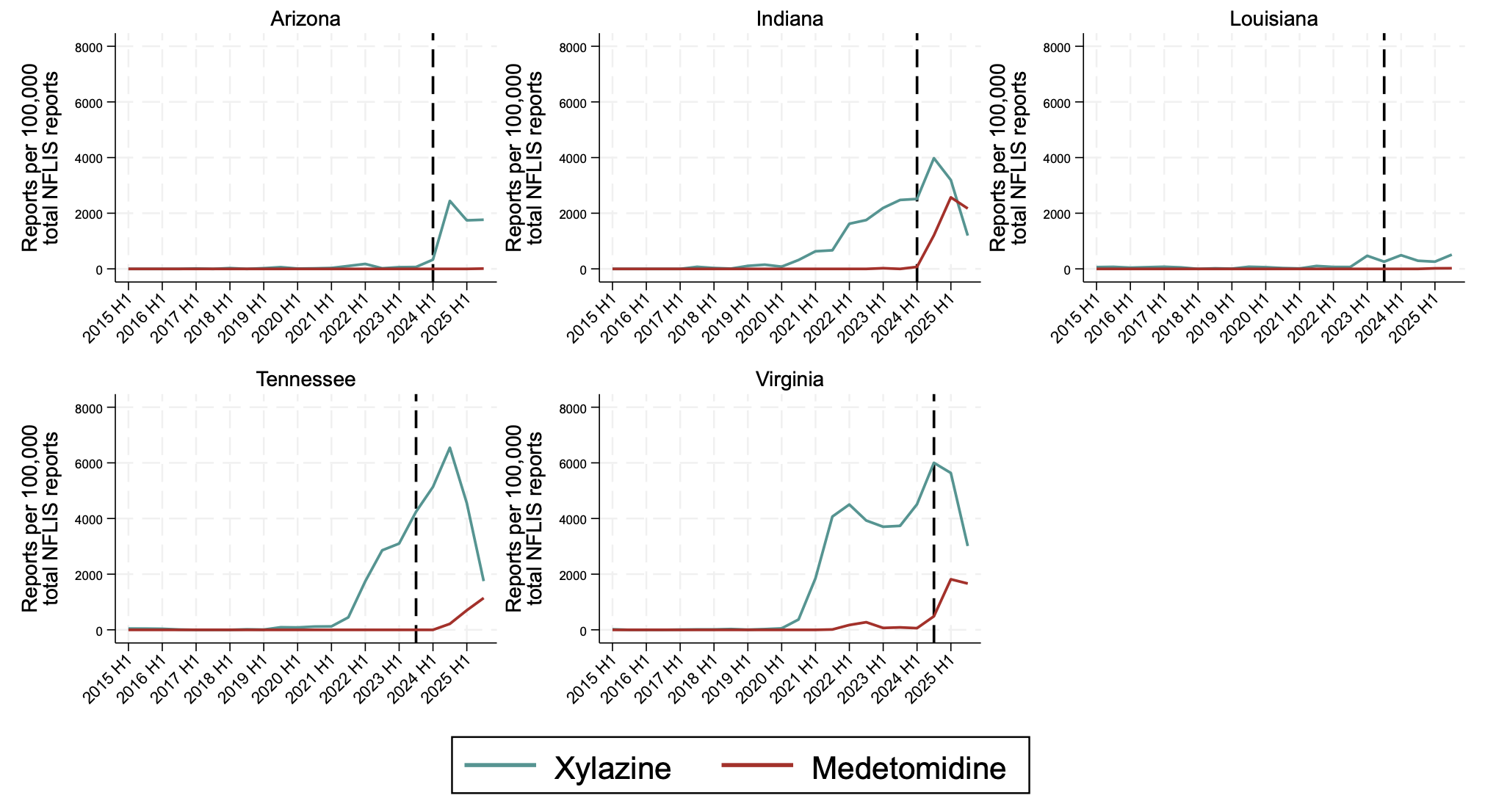

**Caption.** State-level trends in xylazine and medetomidine reports per 100,000 total drug reports in the National Forensic Laboratory Information System (NFLIS) among states that enacted policies criminalizing xylazine but did not formally schedule it as a controlled substance. The teal line represents xylazine reports and the red line represents medetomidine reports. Vertical dashed lines indicate the semiannual period in which criminalization policies took effect in each state. NFLIS data were obtained on May 7, 2026.

**eFigure 3. Event-Study Coefficients of Changes in Xylazine and Medetomidine Report Rates Following State Xylazine Scheduling, Excluding Florida**

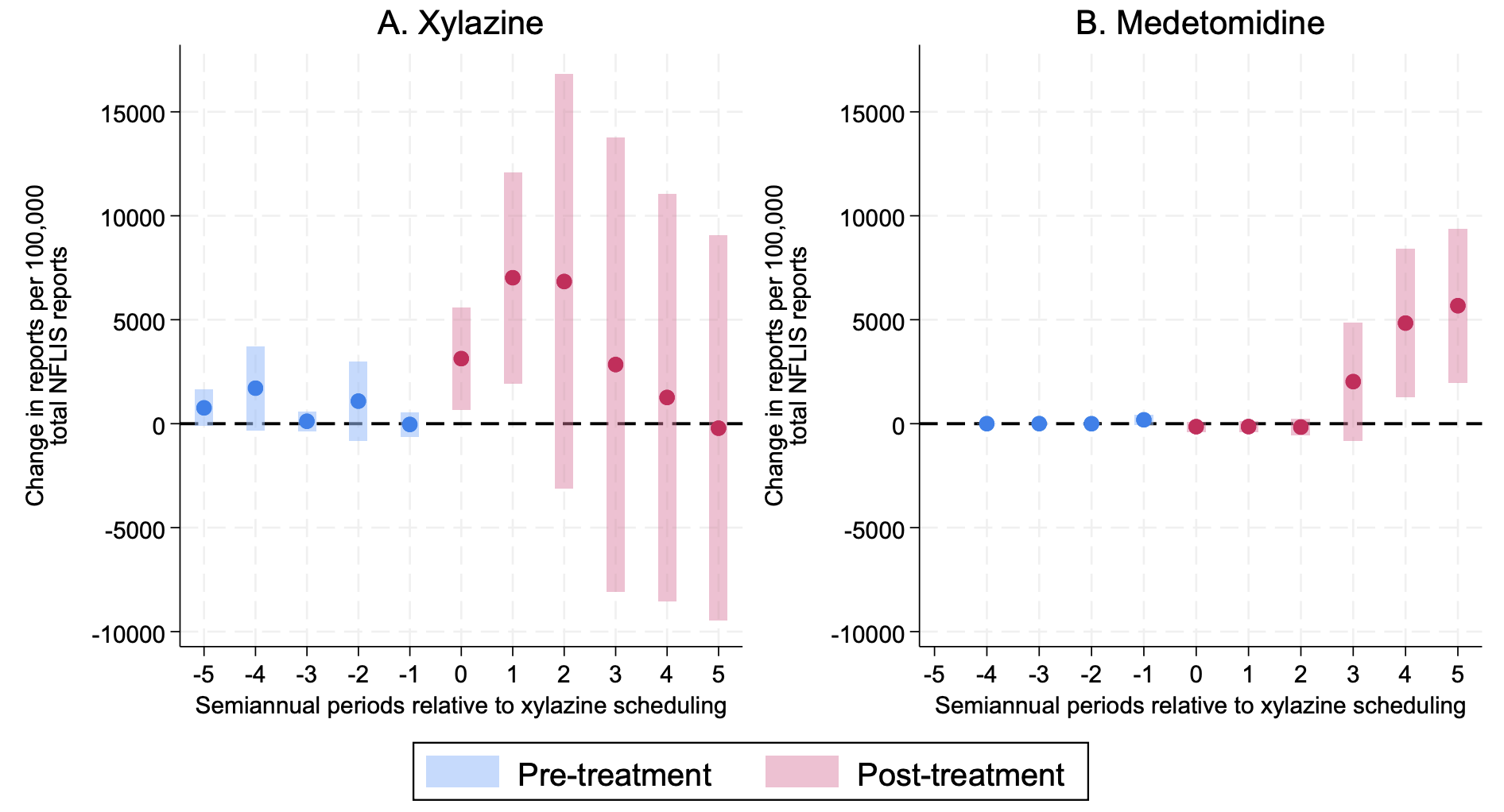

**Caption.** Event-study coefficients of changes in xylazine and medetomidine reports per 100,000 state-level drug reports in the National Forensic Laboratory Information System (NFLIS) following state xylazine scheduling. Points represent estimated treatment effects and shaded bars represent 95% CIs. Blue estimates denote pre-treatment periods and pink estimates denote post-treatment periods. Five semiannual periods before and after scheduling were examined. Massachusetts was excluded because xylazine is covered under its longstanding Schedule VI framework. Florida was excluded because its xylazine scheduling occurred substantially earlier than scheduling in other states. Arizona, Indiana, Louisiana, Tennessee, and Virginia were excluded because they enacted regulations criminalizing xylazine but did not formally schedule it. NFLIS data were obtained on May 7, 2026.

**eFigure 4. Event-Study Coefficients of Changes in Xylazine and Medetomidine Report Rates Following State Xylazine Scheduling or Criminalization**

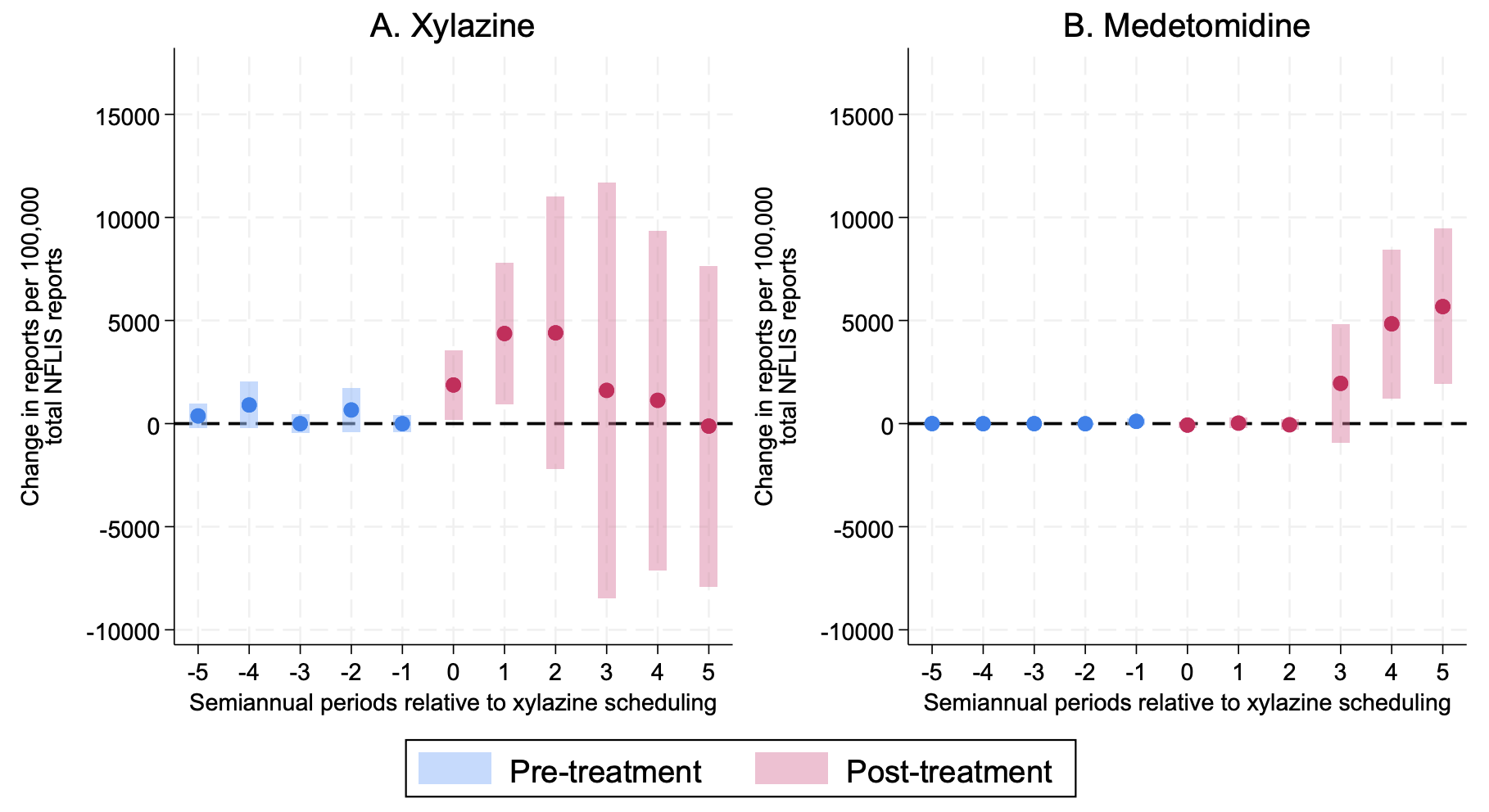

**Caption.** Event-study coefficients of changes in xylazine and medetomidine reports per 100,000 state-level drug reports in the National Forensic Laboratory Information System (NFLIS) following state xylazine scheduling. Points represent estimated treatment effects and shaded bars represent 95% CIs. Blue estimates denote pre-treatment periods and pink estimates denote post-treatment periods. Five semiannual periods before and after scheduling were examined. Massachusetts was excluded because xylazine is covered under its longstanding Schedule VI framework. Arizona, Indiana, Louisiana, Tennessee, and Virginia were added to the treatment group because they enacted regulations criminalizing xylazine, despite not formally scheduling it. NFLIS data were obtained on May 7, 2026.
